# Plasma proteomics reveals clinical and mechanistic heterogeneity among individuals who develop coronary artery disease

**DOI:** 10.64898/2026.06.10.26355410

**Authors:** Yefeng Yang, Dandan Tan, Julia Carrasco-Zanini-Sanchez, Chen-Yang Su, Sirui Zhou, Satoshi Koyama, Pradeep Natarajan, Claudia Langenberg, Tianyuan Lu, Satoshi Yoshiji

## Abstract

**BACKGROUND:** Individuals who develop coronary artery disease (CAD) are clinically and mechanistically heterogeneous, and understanding this variation is crucial for precise risk stratification and tailored interventions. However, the molecular mechanisms that connect these two kinds of heterogeneity remain unclear, limiting progress toward biologically grounded risk stratification and targeted interventions. Here, we investigated the heterogeneity of individuals who develop CAD by leveraging plasma proteomic signatures, placed individuals along continuous metabolic gradients and revealed the molecular programs underlying these patterns, thereby linking mechanistic variation to clinical heterogeneity.

**METHODS AND RESULTS:** From 42,803 UK Biobank participants, including 3,713 individuals who developed CAD within 10 years (incident CAD), we first identified a 320-protein panel from 2,923 baseline proteins that improved prediction of incident CAD beyond clinical risk scores. Using reverse graph embedding, we reduced the proteomic data to two dimensions and mapped each incident case onto the resulting two-dimensional latent proteomic space. These proteomic dimensions show significant associations with cardiometabolic and kidney-related clinical markers. The patterns were replicated in the EPIC-Norfolk study. Phenome-wide Cox regression analyses further linked these proteomic dimensions to 10-year incidence rates for various diseases, including type 2 diabetes, obesity, and chronic kidney disease (CKD). Furthermore, adding the proteomic dimensions to clinical variable-based Cox regression model improved prediction of 10-year incidence of CKD and other diseases, demonstrating the value of proteomic dimensions beyond conventional clinical risk factors. Moreover, individuals with prevalent CAD (diagnosed before proteomic sampling) exhibited high, metabolically adverse dimension values, indicating that these axes capture cumulative metabolic burden. Pathway enrichment analyses implicated altered extracellular matrix organization and immune programs among the proteins contributing to the proteomic dimensions.

**CONCLUSIONS:** Our findings demonstrate that plasma proteomic signatures can dissect the heterogeneity of individuals who develop CAD in continuous phenotypic gradients, improve prediction of CAD and comorbidities, and map underlying biological mechanisms.

**Clinical Perspective:** *What is New?:*

- In 42,803 UK Biobank participants, baseline plasma proteomics identified a protein panel that improved prediction of incident coronary artery disease (CAD) beyond conventional clinical risk scores, including AHA PREVENT, and defined two continuous proteomic dimensions that captured clinically relevant heterogeneity among individuals who later developed coronary artery disease; these patterns were replicated in EPIC-Norfolk cohort.
- These proteomic dimensions captured distinct cardiometabolic and kidney-related patterns, improved prediction of multiple comorbidities, including chronic kidney disease, beyond clinical risk factors, and reflected biological programs involving metabolic, immune, and extracellular matrix pathways.

*What Are the Clinical Implications?:*

- Plasma proteomics can serve as a biologically grounded tool to improve prediction of CAD and related comorbidities, characterize clinically relevant heterogeneity, and provide disease-relevant information beyond standard clinical risk-factor measurements.

## Introduction

Coronary artery disease (CAD) remains a leading cause of morbidity and mortality worldwide^1^ and is characterized by complex pathophysiological mechanisms^2^. There are substantial inter-individual differences in clinical phenotypes and comorbidity profiles among individuals who develop CAD, which should be considered for an effective personalized approach to the prevention of CAD and related comorbidities^3–6^.

Two major data sources are commonly used to dissect CAD heterogeneity: genetic association studies and clinical phenogrouping. On the genetic side, recent large-scale genome-wide association analyses of CAD, including meta-analyses exceeding one million participants, have identified hundreds of risk loci and implicated pathways spanning lipid metabolism, vascular biology, and inflammation, providing strong population-level clues about causal biology^7,8^. Polygenic scores can also stratify inherited susceptibility (including CAD)^9,10^, but genetics alone typically explains only part of inter-individual variability because CAD phenotypes are strongly shaped by non-genetic influences such as lifestyle, comorbidities, and treatment^11–13^. Conversely, recent studies have highlighted CAD heterogeneity primarily using clinical variable-based phenogrouping to define subtypes of CAD and related cardiometabolic diseases^14–16^. For instance, unsupervised machine learning approaches have identified phenotypically distinct CAD subtypes with divergent clinical trajectories^16^. These clusters offer a practical way to summarize individual-level CAD heterogeneity for risk stratification, although whether discrete cluster labels add information beyond clinical variables remains unclear^17^. In addition, efforts to connect clinically defined heterogeneity to underlying genetic drivers or molecular mechanisms have shown only limited success^18^. Integrating intermediate molecular layers between genome and phenome, such as transcriptomics, epigenomics, metabolomics, and proteomics—offers a more direct bridge from genotype to clinical variables and may enable more actionable, biology-grounded stratification for precision medicine^19,20^.

Plasma proteins represent a promising avenue in this regard. They are a central layer in the molecular cascade from gene to phenotype. As key regulators and effectors of pathological processes, plasma proteins play a critical role in CAD pathophysiology^21,22^. Compared with clinical variables, plasma proteins may more directly capture underlying molecular and biological changes, as they are more proximate to genetic regulation and often serve as direct effectors of disease processes ^23,24^. While the genome provides greater stability, its inherent distance from the phenome sometimes limits its power for individual-level CAD classification, as indicated by the substantial overlap in CAD polygenic risk scores between cases and controls^25^. In contrast, the plasma proteome, situated centrally within the multi-omics cascade between the genome and phenome, represents a valuable candidate for identifying disease subtypes at the individual-patient level with a biological underpinning. Advances in high-throughput proteomics now enable large-scale quantification of circulating proteins^26–28^, providing new opportunities to investigate dynamic, disease-relevant biological processes^29,30^ and supporting more precise risk assessment^31,32^ and targeted therapeutic interventions.

Based on these considerations, we hypothesized that plasma proteomic heterogeneity could serve as a bridge between clinical heterogeneity and underlying biological mechanisms in individuals who develop CAD. To test this hypothesis, we focused our analyses on three questions: First, can baseline plasma proteomic variation reveal structured heterogeneity among individuals who develop CAD? Second, does this proteomic heterogeneity capture differences in clinical profiles, comorbidity risk, and the disease trajectory of CAD? Third, what molecular processes are captured by this proteomic heterogeneity, as revealed by pathway enrichment? Through this study, we sought to determine whether plasma proteomics provides an interpretable, biologically grounded representation of heterogeneity among individuals who develop CAD.

## Methods

### Ethics

Both the UK Biobank and European Prospective Investigation into Cancer (EPIC)-Norfolk studies obtained approval from their respective institutional review boards. The present analysis was approved by the UK Biobank (application no. 73958) and the Norfolk Research Ethics Committee (no. 05/Q0101/191). All participants provided written informed consent.

### UK Biobank data

The UK Biobank is a large-scale prospective study comprising nearly half a million volunteers recruited from across the United Kingdom between 2006 and 2010^33^. Participants were aged 40–69 at enrollment, underwent various assessments, completed lifestyle and medical questionnaires, and provided biological samples. Incident events were tracked through links to electronic health records.

The UK Biobank Pharma Proteomics Project (UKB-PPP) performed large-scale proteomic analyses on blood plasma samples from 54,219 UK Biobank participants using the antibody-based Olink Explore 3072 Proximity Extension Assay (PEA) platform, measuring 2,923 proteins. This set included 46,595 individuals randomly selected at baseline, 6,376 participants chosen by members of the UKB-PPP consortium, and 1,268 individuals enrolled in the COVID-19 repeat-imaging study^34^. For this study, we focused on a cohort within the UKB-PPP to examine the heterogeneity of individuals who develop CAD through proteomic data.

### Clinical variables information

Information on baseline clinical variables was obtained from UK Biobank health questionnaires, which included: age at baseline (data field ID 21022), sex (data field ID 31), measured body mass index (BMI, data field ID 21001), body fat percentage (BFP, data field ID 23099), hemoglobin A1c (HbA1c; data field ID 30750), triglycerides (TRIG, data field ID 30870), high-density lipoprotein cholesterol (HDL, data field ID 30760), creatinine (CREAT, data field ID 30700), low-density lipoprotein cholesterol (LDL; data field ID 30780), systolic blood pressure (SBP; data field ID 4080) and alanine aminotransferase (ALT, data field ID 30620).

### Definition of incident CAD cases

Incident CAD cases were defined as individuals that developed CAD within 10 years after protein measurements. We defined the baseline as the date for protein sample collection. We integrated three sources from the UK Biobank: first, ICD-10 hospital records were queried for diagnoses, and the earliest date of any such code was recorded per participant. Second, operation records were screened for OPCS-4 codes indicative of coronary procedures. Third, death records were checked as a cause of death. The earliest date among these three data sources was designated as the CAD event date for each CAD case, provided it occurred after the date of protein sample collection. Additionally, individuals with any self-reported, physician-made diagnosis of CAD at the baseline visit, or with prevalent CAD before sample collection were excluded. Details of the relevant codes are listed in **Table S1**^35^.

### Proteomic data processing

In the UKB-PPP, 2,923 distinct proteins were measured from EDTA plasma samples across eight protein panels (cardiometabolic, cardiometabolic II, inflammation, inflammation II, neurology, neurology II, oncology, and oncology II). Raw counts were normalized to yield NPX (Normalized Protein eXpression) values on a log₂ scale.

In our analysis, we first used the R package missForest (version 1.5)^36^ to impute missing protein values. We excluded participants with more than 50% missing protein measurements, proteins with more than 50% missing measurements across participants, and participants lacking age or sex information. We performed imputations separately for each of the eight protein panels, incorporating age and sex as covariates. After imputation, we removed the participants without testing center, batch, time for Olink processing, and ten genetic principal components (covariates). We then applied rank-based inverse-normal transformation to the proteins and regressed out the effects of age, sex, and the aforementioned covariates. Finally, we re-applied rank-based inverse-normal transformation to the resulting residualized protein values, yielding standardized protein values for subsequent protein selection and analyses to capture variance in plasma protein levels that are not primarily driven by age, sex, or ancestry. Hereafter, we refer to these residualized and transformed values as “standardized protein levels”, which we used in the subsequent analyses. Information of 2,923 proteins is available in **Table S2**.

## Step 1: Protein selection

We utilized least absolute shrinkage and selection operator (LASSO) regression to identify the most predictive proteins for incident CAD (Hereafter, the identified proteins refer to CAD-related proteins). Here, proteins refer to each protein’s standardized protein level. We split the dataset into two subsets, using 70% for model development and 30% for evaluation. In the model development section, we included all proteins as predictors and incident CAD as the outcome, employing 5-fold cross-validation to tune each model’s hyperparameters. Proteins that had nonzero coefficients were selected (CAD-related proteins). In the evaluation section, we evaluated whether the CAD-related proteins could improve predictions of incident CAD beyond three established clinical risk scores: QRISK3, Pooled Cohort Equations (PCE) and the Predicting Risk of Cardiovascular Disease EVENTs (PREVENT) equations. QRISK3 is a well-established cardiovascular risk prediction model that calculates an individual’s 10-year risk of developing cardiovascular disease. It incorporates a broad range of clinical variables, including age, sex, blood pressure, cholesterol levels, smoking status, diabetes, and other comorbid conditions^37^. Similarly, the PCE risk score estimates the 10-year risk of a major atherosclerotic cardiovascular disease (ASCVD) event, based on factors such as age, sex, total and HDL cholesterol, systolic blood pressure, smoking status, and diabetes^38^. As for PREVENT, it is a newer American Heart Association risk equation that estimates 10-year (and 30-year) risk for total cardiovascular disease and provides outcome-specific estimates for ASCVD and heart failure, using cardiovascular–kidney–metabolic health factors including age, sex, systolic blood pressure and antihypertensive treatment, lipid measures, current smoking, diabetes, statin use, kidney function (estimated glomerular filtration rate, eGFR), and BMI, with optional refinement using hemoglobin A1c (HbA1c), urine albumin-to-creatinine ratio (UACR), and a social deprivation index^39^.

To assess the performance, we built six cox proportional hazards models, each using incident CAD as the outcome: three relied solely on QRISK3, PCE or PREVENT, while three combined either QRISK3, PCE or PREVENT with the model-derived probabilities from the CAD-related proteins. We then compared model performance using the Harrell’s C (C-index).

## Step 2: Bridging clinical and mechanistic heterogeneity through plasma proteomics

### DDRTree

DDRTree enables both dimensionality reduction and principal tree construction through reversed graph embedding.^40,41^. It has been previously applied to study the heterogeneity of type 2 diabetes (T2D) using T2D-related phenotypes^14^. In our study, we used the DDRTree implementation from the Monocle2 package (version 2.32.0)^42^ to reduce the dimensionality of high-dimensional protein data. This method generates a principal tree with multiple branches, where individuals within the same branch or nearby principal trunk tend to exhibit similar protein characteristics.

We applied DDRTree to incident CAD cases, using CAD-related standardized protein values as input. The method produced two reduced dimensions (proteomic dimensions, dimension 1 and dimension 2) for these cases. We then used the state function in Monocle2 to assign clusters based on the proteomic dimensions, manually merging adjacent clusters containing fewer than 100 individuals. To explore the distribution patterns of clinical variables across the tree, we visualized incident CAD cases by plotting their rank-based normalized clinical variables against the two DDRTree dimensions. The axes were oriented to ensure an increasing trend for BMI.

### Association analysis with dimension values

To examine if the dimension values were indicative of clinical risk, we regressed each of the nine clinical variables (BMI, body fat percentage, HbA1c, ALT, creatinine, triglycerides, SBP, HDL, and LDL, hereafter we refer them to CAD-related clinical variables) on proteomic dimensions (dimension 1 and dimension 2), respectively.

### Phenome-Wide Association Studies (PheWAS)

We conducted a PheWAS to identify diseases (329 in total) strongly associated with dimension 1 or dimension 2. Cox models were built using each disease as the outcome and dimension 1 or dimension 2 as the predictor. Participants were classified as incident cases if they had the relevant ICD-10 code^43^, with their earliest recorded disease diagnosis date used for analysis. Time-to-event was calculated as the difference between the date of protein collection and the earliest disease diagnosis date. For controls, time-to-event was defined as the time from protein collection to the earliest of the following: date of death, date of loss to follow-up, or 10 years after sample collection. Diseases were considered statistically significant associated with proteomic dimensions if their false discovery rate (FDR)-adjusted p-value was below 0.01.

### eGFR trajectory analysis

We estimated individual estimated glomerular filtration rate (eGFR) slopes from serum creatinine measured at two visits (data field ID 30700) and the corresponding ages at assessment (data field ID 21003). To improve statistical power beyond the 104 incident CAD cases with measurements, we analyzed all 1,462 participants with proteomic data and two creatinine measures and projected them onto the two-dimensional proteomic space using the mapping function (**Supplementary Note 1)**. We then fit linear regression models with eGFR slope as the outcome and proteomic dimension values as predictors, adjusting for baseline age and baseline eGFR.

### Incremental value of proteomic dimensions beyond CAD-related clinical variables

To assess whether the proteomic dimensions captured more information beyond nine CAD-related clinical variables, we compared, for each 329 disease outcome in PheWAS, a baseline Cox model including nine CAD-related clinical variables (BMI, body fat percentage, HbA1c, ALT, creatinine, triglycerides, SBP, HDL, and LDL) with an extended model that additionally included dimension 1 and dimension 2. Incremental discrimination was quantified using Harrell’s C-index, and the change in C-index (ΔC) was calculated as the difference between the extended and baseline models. We also evaluated category-free net reclassification improvement (NRI) at 10 years using the nricens R package, comparing predicted risks from the two models. Positive ΔC and NRI values indicate improved prediction after inclusion of the proteomic dimensions. For visualization, diseases were ranked by ΔC or NRI in each disease categories defined in PheWAS, and the top 50 diseases with highest C-index improvement are shown.

### Validation analysis in EPIC-Norfolk cohort

To provide evidence of generalizability, we tested the association of the proteomic dimensions and nine CAD-related clinical variables in the European Prospective Investigation into Cancer (EPIC)-Norfolk study^44^. The EPIC-Norfolk study is a cohort of 25,639 middle-aged individuals from the general population of Norfolk, a county in Eastern England. The study was approved by the Norfolk Research Ethics Committee (ref. 05/Q0101/191). Participants provided written informed consent. Serum samples from the baseline assessment (1993 - 1997) that had been stored in liquid nitrogen were used for proteomic profiling of a randomly selected subcohort (N = 749) and a Type 2 Diabetes (T2D) case-cohort study (N = 1173), using the Olink® Explore 1536 and Olink® Explore Expansion panels, targeting 2923 unique proteins by 2941 assays.

Participants from the EPIC-Norfolk study were flagged for mortality at the UK Office of National Statistics and vital status was ascertained for the entire cohort. Death certificates, hospitalisation data and cancer registry data were obtained using National Health Service numbers through linkage with the NHS digital database. Electronic health records were coded by trained nosologists according to the International Statistical Classification of Diseases and Related Health Problems, 9th (ICD-9) or 10th Revision (ICD-10). CAD was defined through ICD-9 codes 410-414 and ICD-10 codes I20-I25 if they were registered on the death certificate (as the underlying cause of death or as a contributing factor) or as the cause of hospitalization. Given the long-term follow-up of EPIC-Norfolk included the ICD-9 and ICD-10 coding system, codes were consolidated. We mapped the CAD cases (N = 233) onto the low-dimensional space trained on the UK Biobank data using the mapping function (**Supplementary Note 1**) and visualized their distribution. To investigate potential associations, we performed linear regression analyses to study the associations between these dimensions and the CAD-related clinical variables.

### Pearson-Correlation comparison of top CAD related-proteins to all CAD-related proteins

For each group of CAD-related proteins (top 10, 20, 30, 40, and 50 ranked by their coefficients from the LASSO model), we first obtained the associations between dimension 1 or dimension 2 and nine CAD-related clinical variables. We then concatenated the nine coefficients into a vector and computed its two-sided Pearson correlation with the corresponding associations derived from all 320 CAD-related proteins for each proteomic dimension. Groups with correlation ≥ 0.79 (*P* < 0.001) were considered to exhibit a similar trend to the full set.

### Proteomic contributors to each dimension value

We extracted the weight matrix (W) from the DDRTree analysis, which indicates each protein’s absolute contribution to the dimension values (**Table S3**). We then ranked the proteins by their LASSO coefficients and identified the top 20 CAD-related proteins for further investigation. To quantify the relative contributions of metabolic and renal signals in dimension 1 and dimension 2, using all 42,803 participants (incident CAD cases and controls), we regressed the standardized value of each of these proteins on BMI and serum creatinine simultaneously. We used the standardized effect sizes (Z statistics = β/SE(β)) to quantify the strength of associations. Stronger metabolic signals in a given proteomic dimension should be reflected by higher Z statistics for BMI, particularly for proteins that contribute more strongly to that proteomic dimension. Similarly, stronger renal signals should be indicated by higher Z statistics for serum creatinine.

### Prevalent CAD comparison

We compared the distribution of individuals with prevalent CAD and incident CAD cases in the DDRTree-derived tree plot. Using our mapping function (**Supplementary Note 1**), we assigned proteomic dimension values to individuals with prevalent CAD by using their CAD-related standardized protein values, allowing us to position them within the space defined by the two proteomic dimension values. We then compared density distributions between prevalent and incident CAD cases on the DDRTree plots.

### Enrichment analysis of the CAD-related proteins

To interpret the CAD-related protein set, we conducted Reactome pathway over-representation analysis using Enrichr (https://maayanlab.cloud/Enrichr/). The input set consisted of proteins selected by LASSO regression. Significance was assessed by FDR-adjusted P values; pathways with FDR < 0.05 were considered significant. For each pathway, we report the overlap (k/M), odds ratio, adjusted P value, and Enrichr combined score (the deviation-from-expectation z-score multiplied by the natural log of the P value) for ranking, with overlapping input genes listed in the “Genes” column.

### Proteomic dimension-resolved pathway between proteomic dimensions

We compared CAD-related protein contributions across the two proteomic dimensions. For each CAD-related protein, we extracted its loading on dimension 1 and dimension 2, then standardized loadings within each dimension using a probit transformation. Specifically, loadings were converted to within-dimension ranks and mapped to standard normal quantiles using the inverse normal cumulative distribution function, yielding probit-standardized loadings. We defined a per-protein contrast as the probit-standardized loading in dimension 2 minus the probit-standardized loading in dimension 1. Proteins were mapped to gene symbols and tested against curated Reactome gene sets. To reduce multiple testing and ensure adequate coverage, we restricted analyses to pathways that were significant in the overall enrichment screen at a false discovery rate below 0.05 and that contained at least ten measured proteins.

Directional pathway enrichment was assessed using a top-k leading-edge permutation test. Within each pathway, proteins were ranked by the absolute magnitude of their contrast values, and the leading edge was defined as thirty percent of pathway members, constrained to a minimum of five and a maximum of thirty proteins. We summarized pathway direction and strength using the signed sum of the leading-edge contrast values. Significance was evaluated by comparing this statistic to a two-sided permutation null generated from five thousand random draws of the same number of proteins sampled without replacement from the full set of measured proteins. Empirical P values were adjusted across pathways using the Benjamini–Hochberg procedure, and pathways with a false discovery rate below 0.1 were considered significant. The reported direction (“Dim2 greater than Dim1” or “Dim1 greater than Dim2”) was determined by whether the signed sum was positive or negative. For visualization, pathways were displayed as bubbles positioned by the negative base-ten logarithm of the false discovery rate, sized by the absolute signed-sum statistic, and colored by direction, with a dashed line indicating the false discovery rate threshold of 0.1.

## Results

### Study overview

Our discovery analysis was based on 42,803 participants (3,713 incident CAD cases and 39,090 controls) from the UK Biobank cohort. Of the 2,923 proteins measured at baseline, 2,920 were detected in more than 50% of participants and were included in the analysis. Sample inclusion criteria and case definition are described in **Supplementary Note 2**. Incident CAD cases were defined as individuals who experienced a CAD event within 10 years of plasma protein measurement, while controls were those who remained free of CAD over the same 10-year period (**Methods; Table S2, S4 & S5**).

We followed a two-step approach (**Figure 1**). In Step 1, as preparation for the next step, we used LASSO regression to identify a subset of baseline proteins relevant to incident CAD. Specifically, we identified proteins that could predict 10-year incidence of CAD. These proteins improved the predictive performance of conventional clinical scores (QRISK3^37^, Pooled Cohort Equations (PCE)^38^, and American Heart Association Predicting Risk of Cardiovascular Disease EVENTs (PREVENT) equations^39^). In Step 2, leveraging these selected baseline proteins, we evaluated each of the three key questions described above.

**Figure 1.**
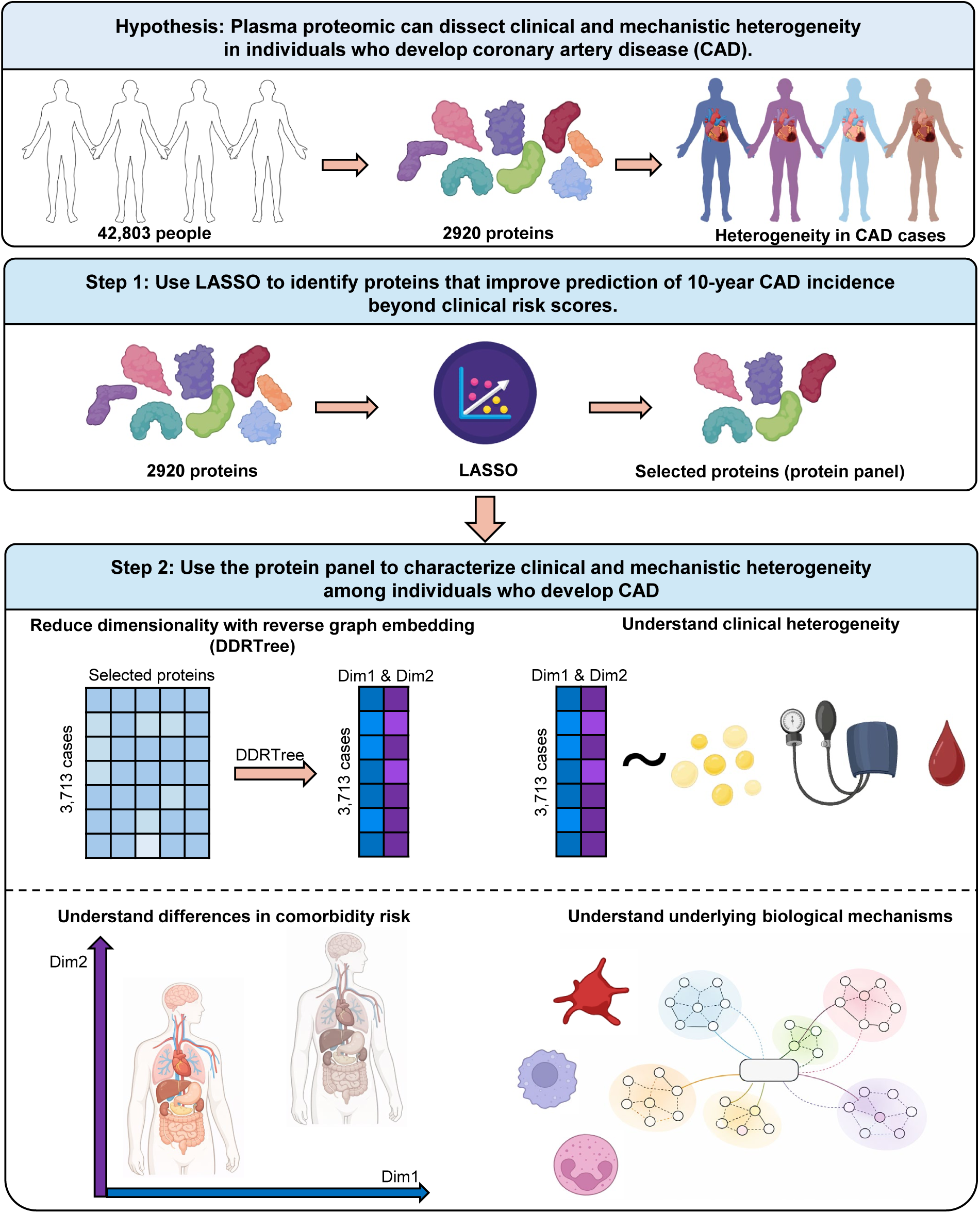
Study hypothesis and design. The study leveraged circulating plasma proteomic signatures to test the hypothesis of plasma proteomics bridge clinical and mechanistic heterogeneity of individuals who develop coronary artery disease (CAD) in a two-step framework. **Step 1.** protein selection (preparatory step): least-absolute-shrinkage-and-selection-operator (LASSO) regression was applied to 2,920 baseline proteins to identify those that predicted 10-year incident CAD; the resulting panel was evaluated for incremental predictive value beyond conventional clinical scores (QRISK3, the Pooled Cohort Equations, PCE and American Heart Association Predicting Risk of Cardiovascular Disease EVENTs (PREVENT) equations). **Step 2.** Bridge clinical and mechanistic heterogeneity of individuals who develop CAD using selected proteins (primary analysis): standardized values of the Step 1 proteins in 3,713 individuals who developed CAD within 10 years (incident CAD cases) underwent dimensionality reduction with DDRTree, yielding two latent proteomic dimensions (Dim1 and Dim2) for each participant. We then performed analyses to test our hypothesis that plasma proteomic heterogeneity could serve as a bridge between clinical heterogeneity and underlying biological mechanisms with the proteomic dimensions.

## Step 1: Protein selection

In this step, we aimed to select a set of proteins relevant to incident CAD as preparation for testing the hypothesis and addressing the three key questions in Step 2, thereby reducing redundancy and increasing computational efficiency. We focused on proteins that predict incident CAD.

### Protein selection process

For each of the 2,920 baseline proteins measured in 3,713 incident CAD cases and 39,090 controls, we first applied rank-based inverse-normal transformation on the baseline protein values and then regressed out covariates (age, sex, and ten genetic principal components) to obtain residualized protein values. These residuals were then applied the rank-based inverse-normal transformation again (mean = 0, SD = 1), as described previously^26,27^. Hereafter, we refer to these residualized and transformed values as “standardized protein levels.” This approach isolates proteomic variance that is not primarily driven by age, sex, or genetic ancestry, thereby enhancing the generalizability of subsequent analyses.

We randomly split the cohort into model development (70%) and evaluation (30%) datasets (**Figure S1a**). Using LASSO, with all proteins as predictors and incident CAD as outcome, we identified a subset of proteins contributing to the prediction of incident CAD, yielding 320 proteins (**Table S6**). In the evaluation dataset, we then used Cox proportional hazards models (Cox models, see **Methods**) to test whether this subset of proteins could enhance CAD prediction in combination with QRISK3^37^, PCE^38^ or PREVENT^39^, all of which rely on traditional clinical risk factors (**Supplementary Note 3**).

### Predictive performance

When only QRISK3, PCE or PREVENT risk scores were used as predictors, the models achieved Harrell’s C (C-index) of 0.747 (95% CI: 0.733–0.760), 0.722 (95% CI: 0.708–0.736), 0.716 (95% CI: 0.700-0.732), respectively (**Figure S1b**). The addition of proteins selected by LASSO improved these scores. Specifically, combining these proteins with QRISK3 increased the C-index (ΔC) by 0.022 to 0.769 (95% CI: 0.764–0.792), combining them with PCE risk score increased the C-index by 0.038 to 0.760 (95% CI: 0.746–0.774), consistent with previous studies^35,45^. The combination of proteomic dimensions with PREVENT risk score also improved the prediction by 0.04 to 0.756 (95% CI: 0.740-0.772). We then used the LASSO-selected proteins for the analysis in Step 2 to test the hypotheses. Hereafter, we refer to these LASSO-selected proteins as CAD-related proteins.

## Step 2: Bridging clinical and mechanistic heterogeneity through plasma proteomics

In this step, we aim to link the clinical and mechanistic heterogeneity of individuals who develop CAD using CAD-related proteins. First, we performed dimensionality reduction on 320 CAD-related proteins using the DDRTree algorithm^41,42^, which yielded two proteomic signature-derived dimensions (dimension 1 and dimension 2) for each individual. These proteomic dimensions enabled projecting participants onto a two-dimensional proteomic space. Second, we related this proteomic space to clinical outcomes and disease trajectories. Third, we investigated the molecular pathways underlying this proteomics-defined heterogeneity. We also performed sensitivity analyses to test whether the proteomic dimensions capture meaningful variation beyond standard clinical risk factors. We report the findings from each stage of the analysis below.

### 1. DDRTree-based dimensionality reduction of CAD-related proteins

First, we asked whether plasma proteomic variation reveals an organized structure of heterogeneity among individuals who develop CAD.

Using the CAD-related proteins in Step 1, we applied Monocle2^41,42^ to reduce the dimensionality of CAD-related standardized protein values in 3,713 incident CAD cases. Monocle2 implements DDRTree to perform dimensionality reduction and principal tree construction, while assigning individuals with similar protein characteristics into branches (i.e. clusters). We selected DDRTree because it captures complex and non-linear structure in the data and arranges individuals along an interpretable tree-like manifold. This method has been successfully used to disentangle the clinical heterogeneity of type 2 diabetes^14^, enabling the identification of both discrete subtypes and continuous gradients of variation. In contrast, principal component analysis (PCA) and Uniform Manifold Approximation and Projection (UMAP) failed to provide discernible clustering patterns among individuals (**Figure S2**). Each incident CAD case was assigned with two proteomic dimension values, dimension 1 and dimension 2, which captured each individual’s overall proteomic profile. We begin by examining the continuous representation.

### 2. Evaluating heterogeneity of individuals who develop CAD via phenotypic spectrum

We asked whether plasma proteomic variation reveals clinical heterogeneity, differences in comorbidity risk, and the disease trajectory of CAD.

We performed our analysis on a continuous phenotypic spectrum, aiming to provide a nuanced evaluation of heterogeneity in CAD-related clinical variables. To explore the distribution patterns of nine CAD-related clinical variables across 3,713 incident CAD cases, we visualized each clinical variable using DDRTree-derived tree plots. These plots revealed distinct patterns in the distribution of standardized CAD-related clinical variables. Specifically, incident CAD cases with high levels of BMI, BFP, ALT, HbA1c, and triglycerides tended to have higher values on both dimension 1 and dimension 2. In contrast, HDL showed the opposite trend, as incident CAD cases with elevated HDL typically had low values on both dimensions. Notably, creatinine (a marker of kidney function) displayed unique patterns, with higher creatinine levels generally corresponding to higher values on dimension 1 and lower values on dimension 2. This contrasted with the patterns of BMI and triglycerides in dimension 2, underscoring the intricate nature of these proteomic dimensions. By comparison, SBP and LDL did not exhibit clear distribution patterns (**Figure 2a**).

**Figure 2.**
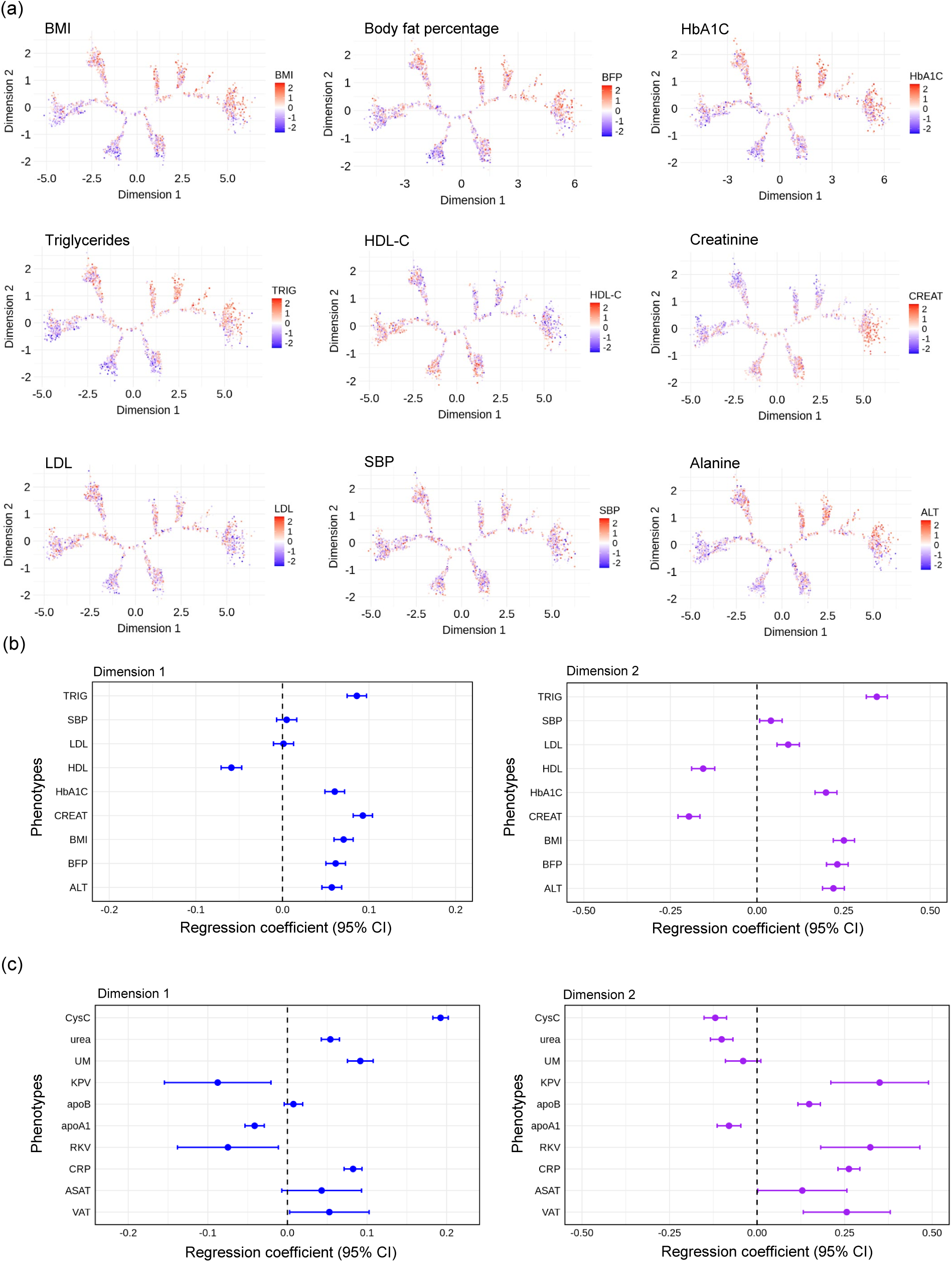
Continuous phenotypic spectrum and heterogeneity of CAD. **a**, The distribution patterns of nine CAD-related clinical variables: body mass index (BMI), body fat percentage (BFP), hemoglobin A1c (HbA1c), alanine aminotransferase (ALT), creatinine (CREAT), triglyceride (TRIG), systolic blood pressure (SBP), high-density lipoprotein cholesterol (HDL) and Low-density lipoprotein (LDL) among 3,713 incident CAD cases across the DDRTree-derived tree plots. **b**, Regression coefficients with 95% confidence intervals, derived from linear regressions assessing the associations between each standardized clinical variable and each unit increase in proteomic dimension (Dimension 1 or Dimension 2). **c**, Regression coefficients with 95% confidence intervals, derived from linear regressions assessing the associations between nine more standardized clinical variable: CysC (Cystatin C), urea (Urea), UM (Urine microalbumin), KPV (Total kidney parenchyma volume), RKV (Right kidney volume), apoA1 (Apolipoprotein A), apoB(Apolipoprotein B), CRP (C-reactive protein), ASAT (Aspartate aminotransferase), VAT(Visceral adipose tissue) and each unit increase in proteomic dimension (Dimension 1 or Dimension 2).

We investigated the linear relationships between nine CAD-related clinical variables and each proteomic dimension (**Methods**). As a result, triglycerides, HbA1c, BMI, BFP, and ALT were positively correlated with both dimension 1 and dimension 2, while HDL showed a negative correlation. Creatinine was positively correlated with dimension 1 but negatively correlated with dimension 2, reflecting distinct renal influences. Meanwhile, LDL and SBP exhibited only marginal positive correlations with dimension 2 **(Figure 2b; Table S7**). These results align with our observation that higher dimension 1 and dimension 2 values tend to coincide with unfavorable metabolic markers (i.e., elevated BMI, BFP, ALT, HbA1c, and triglycerides), whereas higher HDL is associated with lower values on both dimensions. Altogether, this continuous phenotypic perspective highlights complex and distinct patterns of metabolic and renal profiles among incident CAD cases, providing deeper insights into heterogeneity of individuals who develop CAD.

To quantify how much of each proteomic dimension could be explained by the CAD-related clinical variables, we performed variance partitioning using multivariable linear models. We modeled dimension 1 and dimension 2 as functions of nine CAD-related clinical variables. These variables explained a similar proportion of variance in both dimensions (dimension 1, *R*²=0.218; dimension 2, *R*²=0.220), indicating that most dimension variability (∼78%) is not captured by these measured clinical traits. These results support that the proteomic dimensions are related to cardiometabolic or renal risk factors but retain substantial residual structure and capture more information beyond them. We also tested the linear associations between more clinical variables, including Cystatin C, Urea, Urine microalbumin, Total kidney parenchyma volume, Right kidney volume, Apolipoprotein A, Apolipoprotein B, C-reactive protein, Aspartate aminotransferase (AST), Visceral adipose tissue, and each proteomic dimension (**Figure 2c**). Most of these clinical variables show significant associations with either dimension 1 or dimension 2, indicating that those proteomic dimensions capture complex pathophysiological signature. Specifically, dimension 1 showed positive association with Cystatin C (CysC) and negative associations with total kidney parenchyma volume (KPV) and right kidney volume (RKV), whereas dimension 2 showed associations with these kidney-related clinical variables in the opposite direction, consistent with previous finding that dimension 1 and dimension 2 captured distinct renal influences.

Overall, these results suggest that plasma proteomics can dissect the heterogeneity of individuals who develop CAD via continuous phenotypic spectrum.

#### 2.1. Phenome-Wide Association Study (PheWAS)

To further characterize the proteomic dimension values and evaluate whether these proteomic dimensions can predict comorbidity risk, we conducted a PheWAS to examine the associations between diseases and the proteomic dimension values derived from all 42,803 participants (incident CAD cases and controls, we derived the proteomic dimension values of controls by using mapping function, see **Supplementary Note 1)**. Using Cox-regression models, we assessed the relationship between each of the 329 diseases and each proteomic dimension (**Methods, Table S8**).

In **Figure 3**, notably, for both dimension 1 and dimension 2, the top diseases with statistically significant associations were all positively associated with proteomic dimension values (i.e., unfavorable metabolic profiles). The strongest disease associations in both proteomic dimensions involved type 2 diabetes, obesity, and hypertension, all well-established cardiometabolic risk factors closely linked to CAD^46–49^. Non-cardiometabolic diseases were also observed, such as sepsis in each proteomic dimension and sleep apnea in dimension 2 (**Table S9; Figure S3**). Notably, chronic kidney disease, kidney failure and heart failure were significantly associated with dimension 1 but showed no association with dimension 2, which further suggested that dimension 1 captured additional renal signal. These findings indicated that the proteomic dimensions could capture key metabolic profiles associated with cardiometabolic diseases and may also reflect broader systemic disease processes. This reinforces the relevance of the proteomic dimensions in characterizing underlying metabolic dysfunction as well as distinct renal signatures.

**Figure 3.**
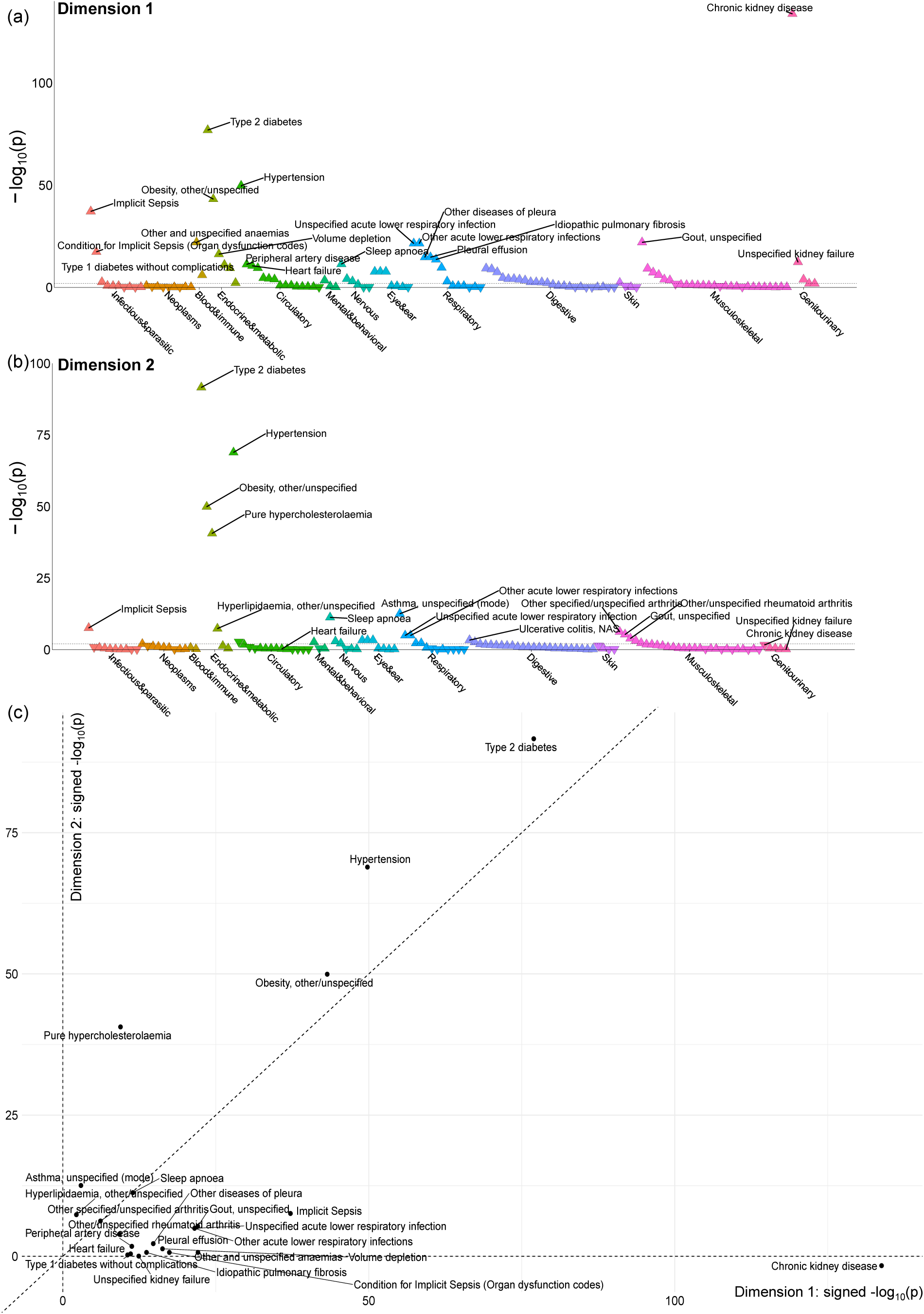
Phenome-Wide Association Study. Cox proportional hazard models were built between each disease and each proteomic dimension from all 42,803 participants. **a**, association for dimension 1, upper triangles indicate a positive association between the proteomic dimension values and disease risk, whereas lower triangles denote a negative association. Diseases were considered statistically significant associated with proteomic dimensions if their FDR-adjusted p-value was below 0.01. **b**, association for dimension 2. **c**, combined comparison of disease associations across the two dimensions. Each point represents a disease endpoint, positioned by its signed −log_10_(P) for Dimension 1 (x-axis) and Dimension 2 (y-axis). The related signs were determined by the direction of associations.

To further characterize the proteomic dimensions with renal function in a clinically interpretable way, we evaluated kidney function by calculating estimated glomerular filtration rate (eGFR) in 1,462 participants with serum creatinine measured at both baseline and follow-up (median follow-up 4 years), using age at each assessment (UK Biobank field 21003) for the two time points. This set included 104 incident CAD cases; proteomic dimension scores were available for all individuals by mapping those without dimension values (n = 1,358) onto the established two-dimensional proteomic space (see **Supplementary Note 1**). Next, we tested whether the proteomic dimensions relate to longitudinal change in kidney function by regressing eGFR slope (mL/min/1.73 m² per year) on each dimension, adjusting for baseline eGFR. Dimension 1 showed a significant negative association with eGFR slope (estimate = −0.083; P = 4.3 × 10⁻^5^), consistent with faster decline at higher dimension 1 values, whereas dimension 2 showed no evidence of association (estimate = 0.067; P = 0.27). These results are reflected in the predicted trajectories, where higher dimension 1 corresponds to a steeper age-related decrease in eGFR (**Figure 4a**), which also supports the notion that dimension 1 captured additional renal signal compared to dimension 2.

**Figure 4.**
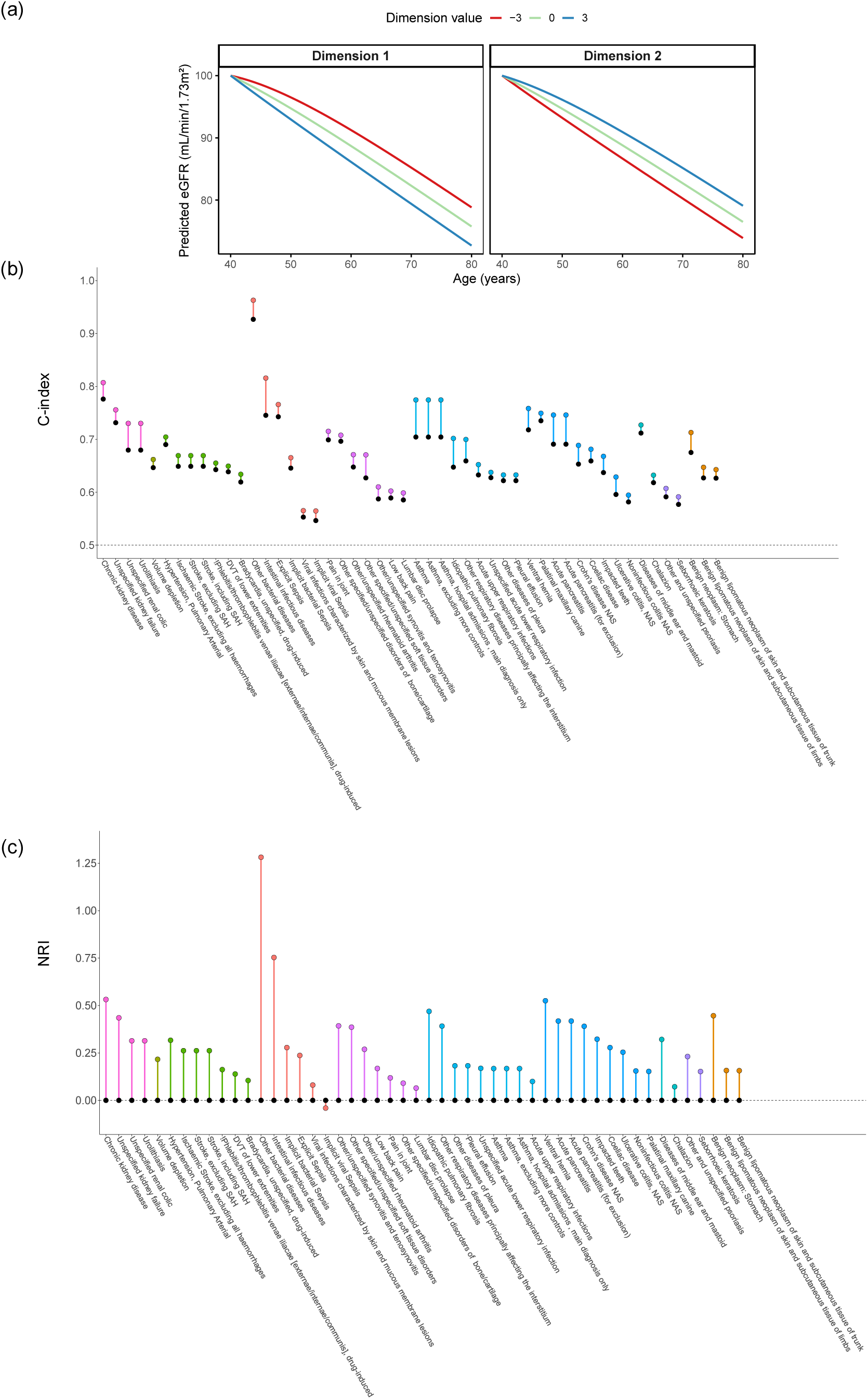
Characterization of proteomic dimensions with renal function via eGFR and improved disease prediction beyond clinical variables. **a**, estimated eGFR is plotted against age separately for dimension 1 (left panel) and dimension 2 (right panel). Lines come from linear models that include an age-by-dimension interaction, evaluated at dimension scores of −3, 0, and +3 (red, green, and blue) while fixing the other dimension at 0. Increasing dimension 1 corresponds to a stronger age-related decrease in eGFR than increasing dimension 2. **b**, comparison of C-index values between baseline Cox model containing the nine CAD-related clinical variables (body mass index (BMI), body fat percentage (BFP), hemoglobin A1c (HbA1c), alanine aminotransferase (ALT), creatinine (CREAT), triglyceride (TRIG), systolic blood pressure (SBP), high-density lipoprotein cholesterol (HDL) and Low-density lipoprotein (LDL)) to an extended model that additionally included proteomic dimensions. Black dots indicate the C-index values for baseline Cox model, while colored dots indicate the C-index values for extended Cox model. We show top 50 diseases with highest C-index improvement. The colors indicate the categories of the diseases defined in Phenome-Wide Association Study. **c**,. Comparison of 10-year category-free net reclassification improvement (NRI) after adding proteomic dimensions to a baseline Cox model including nine CAD-related clinical variables: body mass index (BMI), body fat percentage (BFP), hemoglobin A1c (HbA1c), alanine aminotransferase (ALT), creatinine (CREAT), triglycerides (TRIG), systolic blood pressure (SBP), high-density lipoprotein cholesterol (HDL), and low-density lipoprotein cholesterol (LDL). Black dots represent the null reference value of 0, and colored dots represent the NRI values of the extended model relative to the baseline model. Positive NRI values indicate improved reclassification performance with the addition of proteomic dimensions. The top 50 diseases with the highest C- index improvement are shown. Colors denote disease categories from the Phenome-Wide Association Study.

Furthermore, to test whether the proteomic dimensions capture information beyond the nine CAD-related clinical variables, we conducted model comparisons across the phenome. For each disease, we compared a baseline Cox model including the nine CAD-related clinical variables with an extended model additionally including dimension 1 and dimension 2. Incremental discrimination was quantified as the change in Harrell’s C-index (ΔC), where positive ΔC indicates improved risk ranking after adding the proteomic dimensions. We presented the top 50 diseases with the largest ΔC values. Among these diseases, the median ΔC was 0.020 (range 0.010 to 0.070; **Figure 4b)**. We further evaluated category-free net reclassification improvement (NRI) to assess whether the proteomic dimensions provided nonredundant predictive information beyond the CAD-related clinical variables. Across the top 50 diseases, the median NRI was 0.245, with values ranging from −0.041 to 1.282 (**Figure 4c**), indicating that the addition of the proteomic dimensions generally improved risk reclassification. Notably, among kidney-related diseases, chronic kidney disease and unspecified kidney failure showed ΔC improvements of 0.031 and 0.024, respectively, with corresponding NRI values of 0.532 and 0.435.

#### 2.2. EPIC Norfolk incident cases replicate dimension–phenotype associations

To test portability beyond UK Biobank, we projected 233 incident CAD cases from the independent EPIC-Norfolk cohort^44^ onto the same proteomic space (**Supplementary Note 1**; **Figure S4**). This third evaluation set reproduced the direction of every dimension–phenotype association observed in UK Biobank (**Table S10**) reinforcing the generalizability of the embedding.

#### 2.3. Deriving dimension values from smaller sets of proteins

For future clinical applications, using a smaller, more manageable set of biomarkers can increase feasibility, lower costs, and facilitate implementation. As proof-of-concept exploration, we examined the associations between nine CAD-related clinical variables and the proteomic dimensions derived from the top 10, 20, 30, 40, or 50 CAD-related proteins (ranked by their absolute LASSO coefficients) in 3,713 incident CAD cases. We applied the DDRTree algorithm to generate low-dimensional representations for each subset of proteins in the same manner. Our analysis revealed that the directions of association remained largely consistent across most protein subsets, with only slight variation in creatinine, LDL, and SBP (**Figure 5**, **Table S11**). Moreover, Pearson correlation analysis indicated that using at least the top 20 proteins could maintain trends comparable to those observed with all 320 CAD-related proteins (r = 0.892 for dimension 1, 0.911 for dimension 2). In contrast, patterns for the top 10 proteins showed greater deviation (*r* = 0.775 for dimension 1, 0.279 for dimension 2; **Methods, Table S12**). These findings suggest that a more compact protein panel, consisting of at least the top 20 proteins, may still capture the original signature while maintaining robust predictive capabilities.

**Figure 5.**
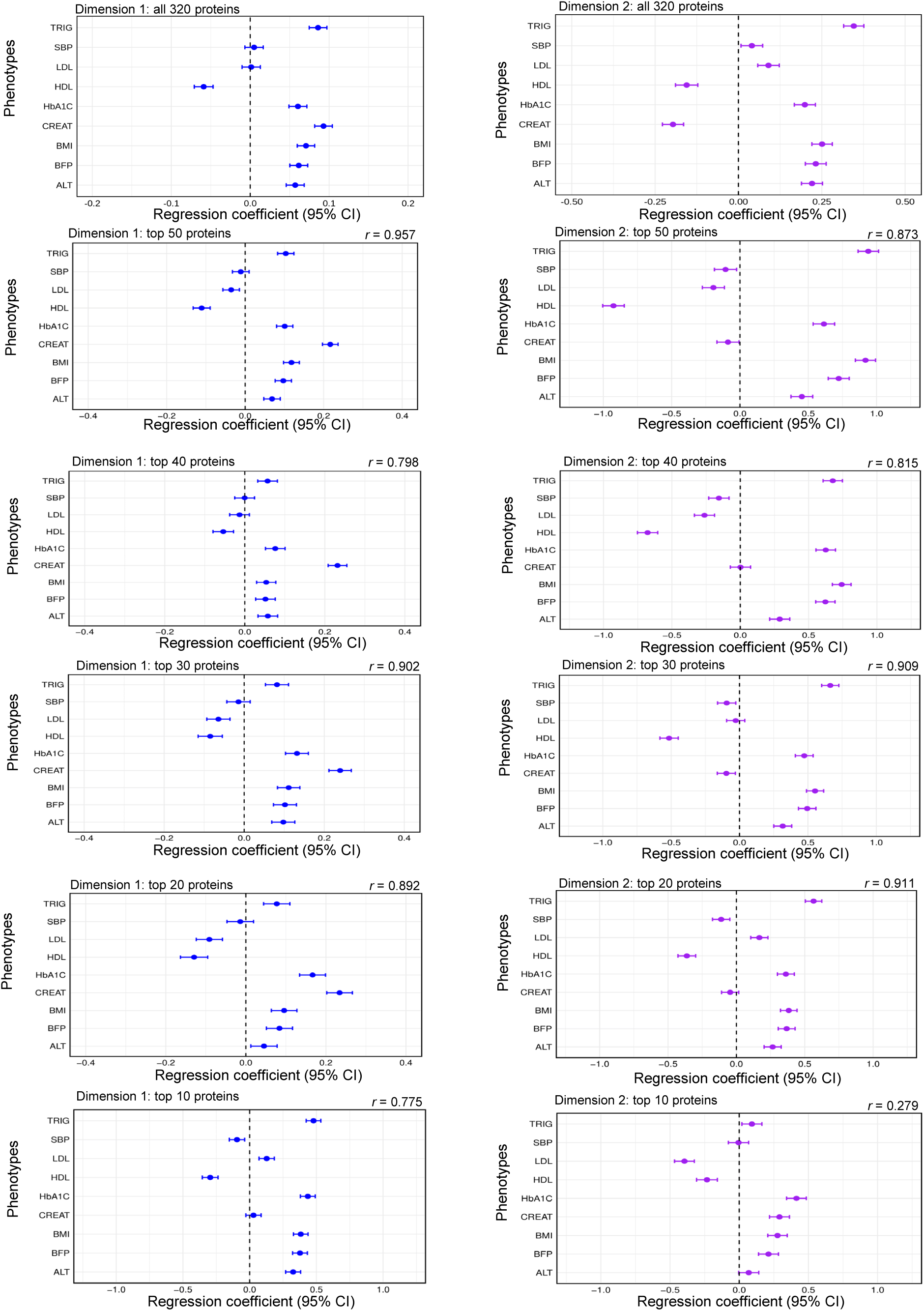
The associations between each clinical variable and each proteomic dimension from top proteins. Regression coefficients with 95% confidence intervals (x-axis) are derived from linear regressions that assess the associations between each clinical variable (rows on the y-axis) and each proteomic dimension (Dimension 1 in blue or Dimension 2 in purple). Each column of panels corresponds to different subsets of top proteins (top 50, 40, 30, 20, or 10), selected by LASSO ranking. The dashed vertical line at zero indicates no effect. Abbreviations for clinical variables include TRIG (triglycerides), SBP (systolic blood pressure), LDL (low-density lipoprotein cholesterol), HDL (high-density lipoprotein cholesterol), HbA1c (hemoglobin A1c), CREAT (creatinine), BMI (body mass index), BFP (body fat percentage), and ALT (alanine aminotransferase). *r* indicates the Pearson correlation coefficients for each subset of top proteins.

#### 2.4. Proteomic contributors to each dimension value

To understand which proteins contribute most to each dimension, we examined the DDRTree weight matrix (W), which quantifies each protein’s absolute contribution to dimensions 1 and 2. (**Table S3**). We observed that these dimensions are defined by a broad combination of proteins, rather than by a small number of strong drivers. We note that some of these proteins may be proxies (i.e., “tagging” underlying effector proteins) because we removed highly correlated proteins during the Step 1 LASSO regression to improve model stability. Moreover, to investigate whether these two proteomic dimensions capture distinct biological axes, we focused on the top 20 CAD-related proteins (ranked by LASSO coefficient) and plotted their dimension-specific weights alongside their relative associations with BMI (representing metabolic status) and creatinine (representing renal function) in **Figure S5**. This bubble-plot analysis suggests that dimension 1 carries a substantial renal (creatinine-related) signal in addition to metabolic influences, whereas dimension 2 is driven primarily by metabolic variation, aligning with the findings from previous sections.

#### 2.5. Proteomic dimensions capture metabolic trajectories of CAD

While the proteomic dimensions capture metabolic dysfunction among incident CAD cases, we hypothesized that they would also stratify individuals with prevalent CAD, i.e. those who developed CAD before proteomic sampling and may therefore reflect established disease (**Table S5**). These 1,331 prevalent cases were excluded from both Step 1 and Step 2 and therefore serve as an independent evaluation set. By using the mapping function (**Supplementary Note 1**), we projected the prevalent cases onto the DDRTree map obtained based on incident CAD cases (**Figure 6**). Prevalent cases were markedly enriched in regions with high dimension 1 and dimension 2 values, which were the most metabolically adverse regions. This aggregation supports the notion that longer CAD duration corresponds to deeper metabolic derangement and illustrates the dimensions’ potential for tracking the severity of underlying metabolic burden.

**Figure 6.**
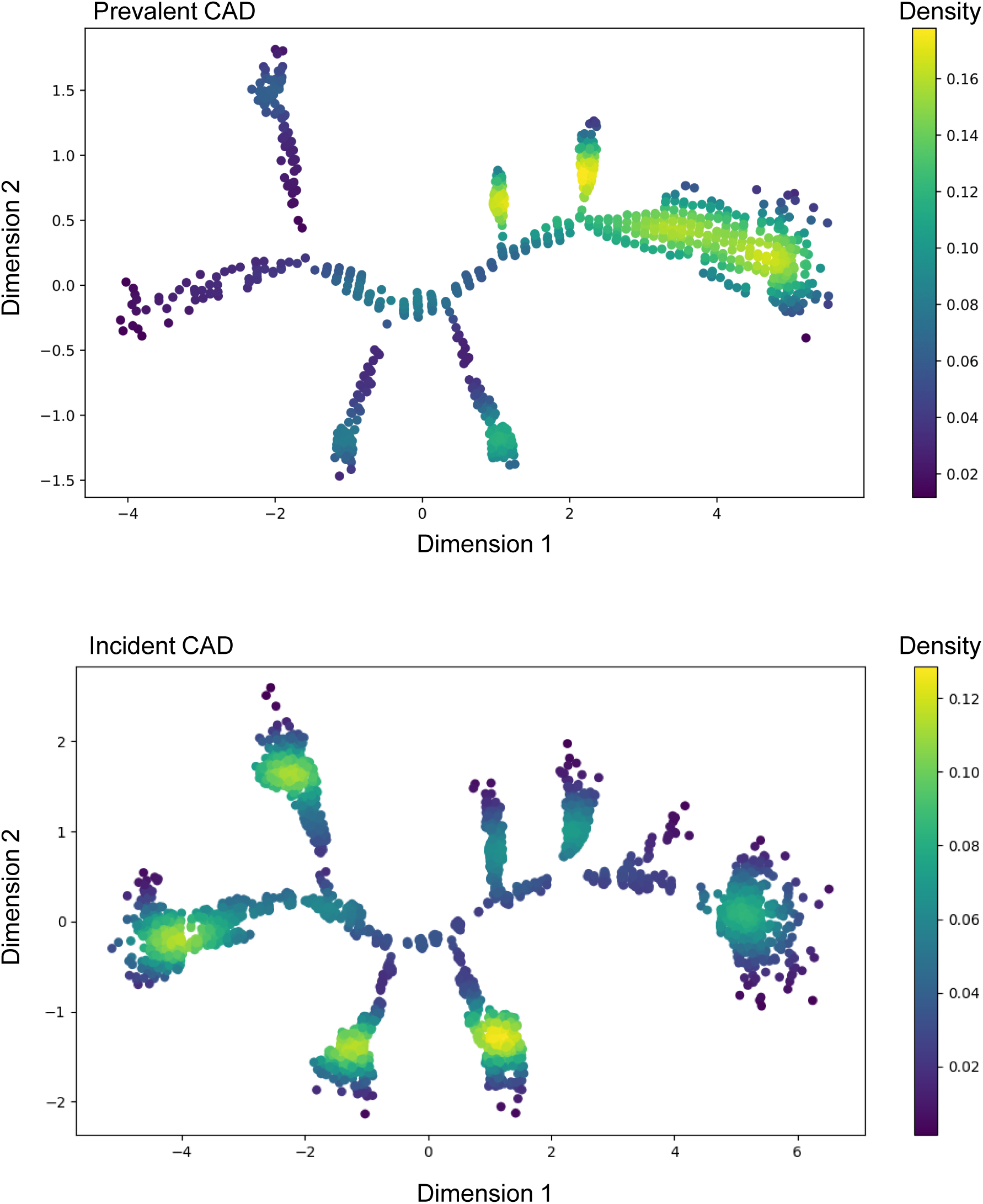
Distribution of individuals with prevalent CAD. The top panel displays the DDRTree-derived dimensional space (Dimension 1 on the x-axis, Dimension 2 on the y-axis) for individuals with prevalent CAD, and the bottom panel shows the corresponding distribution for incident CAD cases. Colors represent density, ranging from low (purple) to high (yellow).

#### 2.6. Proteomic dimensions can define clinically distinct clusters

While our study primarily focused on the continuous dimensions because they can capture finer differences than discrete grouping, in supplementary analysis, we assessed whether the proteomic dimensions could also delineate the discrete clusters. By using Monocle2 (see **Methods**), nine clusters were identified (**Figure S6a**). We investigated the clinical characteristics of each cluster (**Figure S6b**). With low dimension 1 and median dimension 2 values, Cluster 1 comprised individuals who were relatively lean and had higher HDL and lower triglycerides, indicating a favorable metabolic phenotype. With similar dimension 1 values as Cluster 1 but lower dimension 2 values, Cluster 3 and Cluster 4 formed moderate-risk groups with lean body composition, though Cluster 4 exhibited particularly low BMI and triglycerides, coupled with high HDL In contrast, Cluster 8, with similar dimension 2 values as Cluster 1 but higher dimension 1 values, showed the highest average BMI, triglycerides, HbA1c, and ALT, suggesting a “metabolically unhealthy” status. Cluster 9, with similar dimension 2 values as Cluster 8 but higher dimension 1 values, also featured high BMI and low HDL, but also with high creatinine (**Table S13**). We further evaluated whether usage of medication, specifically statin^50^, aspirin, and antihypertensive drug influenced the associations with those clinical variables and found that the overall patterns remained consistent, confirming that the observed heterogeneity reflected underlying biology rather than pharmacological confounding (**Supplementary Note 4**, T**able S14 & S15**, **Figure S7**). The violin plots comparing incident CAD cases with and without myocardial infarction also showed identical distributions for both dimension 1 and dimension 2, indicating that the proteomic dimensions captured pathophysiological variation common to CAD rather than MI-specific processes (**Table S16**, **Figure S8)**.

To assess the risk of future complications captured by the baseline proteomic profile, we calculated the proportion of individuals who experienced incident chronic kidney disease (CKD), heart failure (HF), type 2 diabetes (T2D), obesity, or hypertension which are cardiometabolic comorbidities closely linked to CAD^46–49^, within each cluster over the 10 years after proteomic measurement. We found that hypertension was highest in clusters with elevated BMI and adverse metabolic markers (e.g., Cluster 9). Obesity was likewise most common in Cluster 9, consistent with its higher BMI and body fat percentage. T2D occurred most frequently in these same metabolically adverse clusters, aligning with their raised HbA1c and triglyceride levels (**Figure S9**). CKD incidence was also enriched in Cluster 9, mirroring its elevated creatinine.

These distinct metabolic signatures highlight the heterogeneity of incident CAD and reveal cluster-specific risks for key cardiometabolic diseases. They also demonstrate clinical distinctions among the clusters, supporting the utility of proteomic profiles in refining our understanding of CAD phenotypes and their related cardiometabolic and renal complications.

### 3. Molecular mechanisms captured by proteomic dimensions

Third, we characterized the molecular processes captured by the proteomic dimensions through pathway enrichment and analysis of dimension-specific protein contributions.

#### Reactome pathway enrichment of CAD-related proteins

We ran Reactome pathways analysis^51^ on the 320 CAD-related proteins used to build the proteomic dimensions and identified 36 significant pathways (FDR < 0.05; **Methods**). The enriched terms captured two dominant modules: metabolic and hemostatic processes, including plasma lipoprotein related processes and platelet activation, and immune programs (**Figure 7a**). In addition, extracellular matrix organization was also captured. Overall, these proteins are enriched for coordinated processes spanning metabolic–hemostatic pathways, immune signaling and extracellular matrix organization.

**Figure 7.**
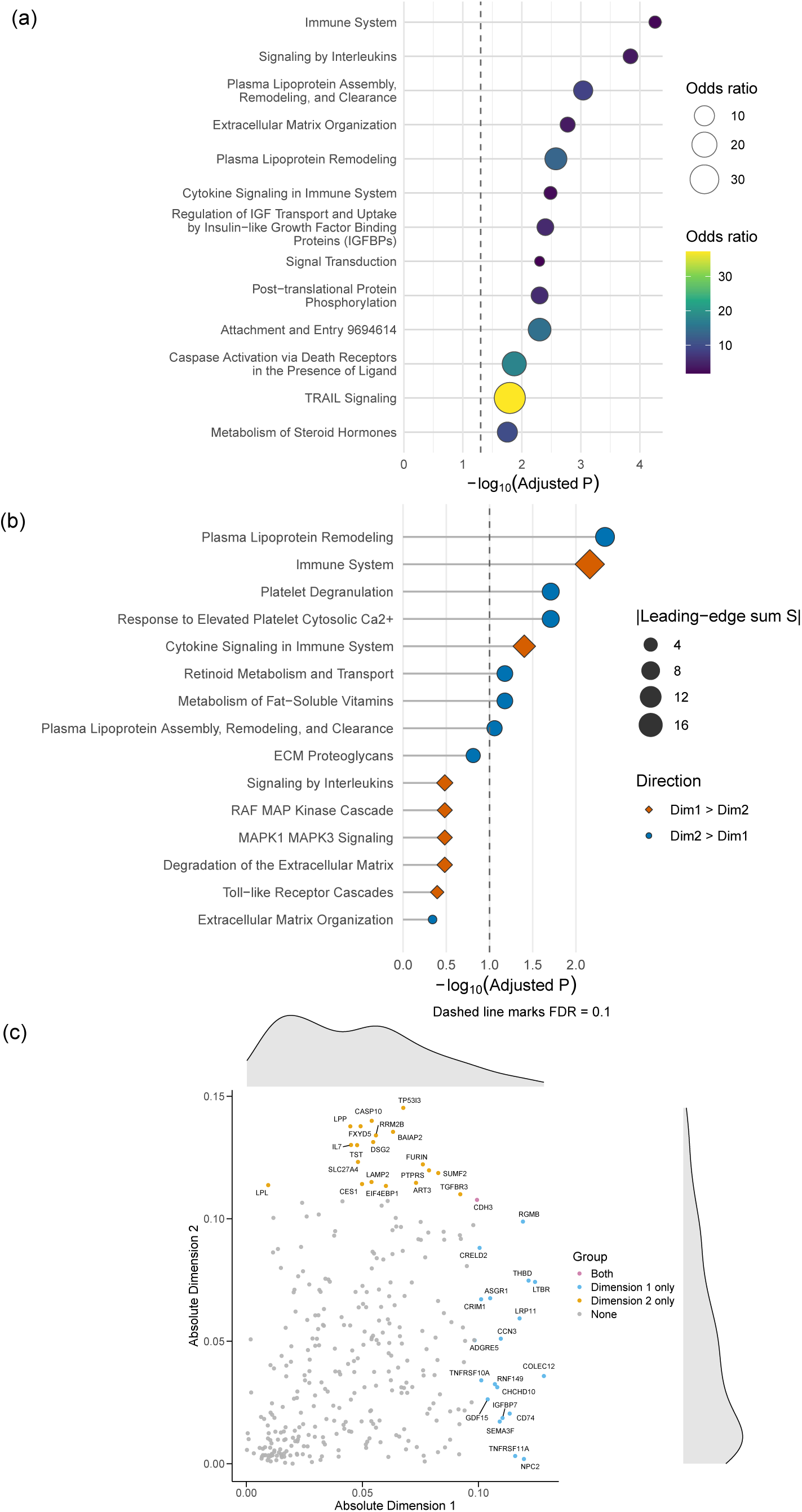
Molecular mechanisms underlying proteomic dimensions. **a**, Reactome enrichment analysis for the 320 proteins selected by LASSO. The dot plot displays the most significantly enriched pathways (FDR < 0.05) ordered by enrichment score; dot size reflects the number of mapped proteins and color represents significance (−log10 FDR). **b**, Differential Reactome enrichment comparing Dimension 2 and Dimension 1 protein weights using a top-k leading-edge permutation approach. Dot size is proportional to the magnitude of the leading-edge statistic (|S|) and point color/shape indicates the predominant direction of the signal (Dim2 > Dim1 vs Dim1 > Dim2); the dashed vertical line denotes FDR = 0.1. We used FDR<0.1 as an exploratory threshold for differential enrichment given reduced power in contrast analysis. **c**, Dimension-specific protein contributors. Each protein is positioned by its absolute loading on Dimension 1 (x-axis) and Dimension 2 (y-axis) according to DDRTree weight matrix (W), with colors indicating whether each protein ranks among the top 20 contributors to Dimension 1 only, Dimension 2 only, both dimensions, or neither.

#### Proteomic dimension-resolved pathway contrasts between dimension 1 and dimension 2

To identify pathways preferentially captured by each dimension, we contrasted per-protein weights between dimension 1 and dimension 2 and applied a top-k leading-edge permutation framework (**Methods, Figure 7b**). Protein weights here refer to the DDRTree weight matrix (W), which captures each protein’s absolute contribution to dimension 1 and dimension 2 (**Table S3**). This analysis indicated that dimension 2 was preferentially enriched for metabolic pathways, including plasma lipoprotein remodeling (P = 2.0 × 10⁻^4^), and platelet-related pathways (P = 3.4 × 10⁻^3^ for platelet degranulation; P = 3.4 × 10⁻^3^ for response to elevated platelet cytosolic Ca2+), whereas dimension 1 showed relatively stronger enrichment for immune pathways (P = 6.0 × 10⁻^4^ for immune system, P = 8.6 × 10⁻^3^ for cytokine signaling in immune system). Thus, while both dimensions capture cardiometabolic biology, they highlight partially distinct modules, with dimension 2 aligning with a metabolic–hemostatic axis and dimension 1 with immune signaling. These differences were underpinned by largely non-overlapping protein contributors across the two proteomic dimensions (**Figure 7c**). The top 20 proteins for each dimension showed almost no overlap, sharing only one protein (CDH3), consistent with distinct molecular drivers.

### 4. Sensitivity analyses: proteomic dimensions beyond clinical risk factors

In this section, we performed sensitivity analyses to further confirm whether the proteomic dimensions could still capture meaningful information beyond nine CAD-related clinical variables, including PheWAS and linear association analyses after regressing out the CAD-related clinical variables.

#### 4.1 Phenome-wide association analysis of clinically adjusted proteomic dimensions after regressing out CAD-related clinical variables

To further test whether the proteomic dimensions capture information beyond CAD-related clinical variables, besides the covariates (age, sex, and ten genetic principal components) we regressed out during Step 1, we also regressed out the nine CAD-related clinical variables and then performed the similar protein selection process and dimension reduction to obtain the new clinically adjusted proteomic dimensions. We repeated the PheWAS (329 diseases) using these clinically adjusted proteomic dimensions. Despite removal of clinical effects, both adjusted proteomic dimensions retained multiple significant disease associations (24 diseases for clinically adjusted dimension 1, 37 diseases for clinically adjusted dimension 2, **Figure S10**), supporting that the proteomic dimensions could reflect residual structure beyond standard clinical measures, indicating the proteomic dimensions are not fully explained by routine clinical phenotyping.

#### 4.2 Clinically adjusted proteomic dimensions retain interpretable biological axes

We further performed linear regression analyses between clinically adjusted proteomic dimensions and more phenotypes to investigate their linear relationships (**Figure S11**). Clinically adjusted dimension 1 retained a renal signature, with higher values significantly associated with higher cystatin C and urinary microalbuminuria and lower kidney and parenchyma volumes, consistent with a residual renal axis. Clinically adjusted dimension 2 aligned with an immunometabolic burden pattern, showing significant positive associations with gamma-glutamyl transferase (GGT) together with higher white blood cell and neutrophil counts. We also tested whether the clinically adjusted proteomic dimensions relate to longitudinal change in kidney function by using the same methods for original proteomic dimensions. Clinically adjusted dimension 1 remained significantly inversely associated with eGFR slope (estimate = −0.097; P = 1.8 × 10⁻^4^), indicating faster eGFR decline at higher clinically adjusted dimension 1 values, whereas clinically adjusted dimension 2 showed no evidence of association (P = 0.65). Overall, these findings indicate that the renal-decline signal is preserved in dimension 1 after clinical adjustment, whereas clinically adjusted dimension 2 still does not capture longitudinal eGFR change.

## Discussion

In this study, we showed that plasma proteomic heterogeneity can bridge clinical and mechanistic heterogeneity in individuals who develop CAD. As key regulators and effectors of pathological processes, plasma proteins also play a critical role in CAD pathophysiology^21,23^. The analyses were designed to answer three questions: (1) Can baseline plasma proteomic variation delineate structured heterogeneity among individuals who develop CAD? (2) Does this proteomic heterogeneity capture differences in clinical profiles, comorbidity risk, and the disease trajectory of CAD? (3) Which molecular pathways and processes underlie this proteomic heterogeneity?

First, we found that baseline plasma proteins could dissect the heterogeneity of individuals who develop CAD through both continuous axes (as well as discrete clusters). Using a LASSO-selected panel of 320 proteins predictive of 10-year incident CAD, DDRTree embedding of 3,713 incident CAD cases generated a two-dimensional proteomic space (dimension 1 and dimension 2), providing a unified representation that captures coordinated molecular variation alongside phenotypic heterogeneity of individuals who develop CAD.

Second, the two proteomic dimensions captured complementary cardiometabolic and renal patterns, leading to distinct comorbidity risk. Both proteomic dimensions tracked adverse metabolic status (higher BMI, HbA1c, ALT, and triglycerides, and lower HDL), but dimension 1 and dimension 2 reflect a different renal signal, supported by their different associations with creatinine, Cystatin C and kidney volumes. In PheWAS analysis, higher values along either dimension were associated with cardiometabolic diseases (type 2 diabetes, obesity, and hypertension), while renal and cardiac complications (chronic kidney disease, kidney failure, and heart failure) were specific to dimension 1, indicating that dimension 1 captured an added renal-risk axis. The longitudinal eGFR analysis reinforced this distinction, with higher dimension 1 predicting faster eGFR decline and no evidence of association for dimension 2. Together, these findings suggest that dimension 1 might reflect a cardiovascular-kidney-metabolic axis, an interplay of metabolic risk factors, renal dysfunction, and cardiovascular pathology that strongly influences morbidity and mortality^52^, whereas dimension 2 more selectively reflected metabolic burden.

Importantly, these proteomic dimensions improved prediction of 10-year incidence for multiple diseases, including CKD: adding the proteomic dimensions to nine CAD-related clinical variables increased Harrell’s C-index by a median of 0.020 (range 0.010 to 0.070) among the top 50 diseases with the greatest C-index improvement, and yielded a median category-free net reclassification improvement (NRI) of 0.245 (range −0.041 to 1.282), supporting added predictive and reclassification value beyond conventional clinical risk factors. Additional sensitivity analyses showed that clinically adjusted dimensions recomputed after removing these clinical effects retained interpretable renal and immunometabolic signatures together with persistent disease associations. Those results suggest that plasma proteomics capture organized variation beyond standard risk factor measurements. In contrast, clustering approaches built solely from established cardiovascular risk factors have been reported to perform comparably to SCORE2, PCE, and PREVENT, with no statistically significant differences in discrimination (C-statistic), suggesting that discrete risk-factor–derived cluster labels may offer limited incremental value beyond conventional clinical models^17^.

To place these axes in a broader disease context, we projected participants beyond the incident-case discovery set into the same proteomic space. Importantly, we observed that individuals with prevalent CAD tended to cluster in the metabolically adverse region of the proteomic dimensional space. This enrichment of prevalent CAD cases in the most metabolically adverse space suggests that a longer disease course corresponds to more advanced metabolic disturbances. In other words, cumulative exposure to risk factors and ongoing disease processes may lead to more pronounced metabolic derangements, which the proteomic dimension values effectively capture. This supports the idea that these dimensions not only reflect phenotypic differences at baseline but may also capture accumulated metabolic burden over time. Projection of incident CAD cases from the independent EPIC-Norfolk cohort reproduced the direction of dimension–phenotype associations observed in UK Biobank, supporting portability of the embedding.

Third, we also explored the molecular pathways underlying these proteomic dimensions. Reactome enrichment analysis of the 320 CAD-related proteins defining the proteomic dimensions highlighted coherent biological modules spanning metabolic and hemostatic programs (including plasma lipoprotein remodeling and platelet-related pathways), immune signaling and extracellular matrix organization. Differential pathway analyses further suggested that dimension 2 preferentially emphasized metabolic–hemostatic pathways, whereas dimension 1 showed relatively stronger immune signal alongside its renal associations, consistent with established roles of lipid, thrombotic and inflammatory programs in CAD^7,18,53,54^. The limited overlap among top contributing proteins supports the interpretation that the two proteomic dimensions capture separable components of CAD biology rather than a single continuum.

Taken together, these results indicate the proteomic dimensions as an intermediate layer linking molecular programs to clinical heterogeneity in individuals who develop CAD. Dimension 2 captures a predominantly metabolic burden axis, whereas dimension 1 reflects overlapping metabolic burden with additional renal features, consistent with its unique associations with CKD risk and eGFR decline. Notably, dimension values derived from as few as 20 proteins largely recaptured the metabolic signatures of the 320-protein dimensions, indicating a practical path toward a compact protein panel that could be deployed more cost-effectively in clinical practice. More importantly, our analyses indicate that these proteomic dimensions are not fully reducible to routine clinical phenotyping: nine CAD-related clinical variables explained only around 22% of the variance in each dimension. Indeed, adding the proteomic dimensions to Cox regression models based on clinical variables improved prediction of 10-year incidence for multiple diseases, including CKD.

Our study has several limitations. First, although individuals of all ancestries were included, the UK Biobank cohort is predominantly European ancestry. Second, despite profiling 2,920 proteins, coverage of some low-abundance or tissue-specific proteins remains incomplete, potentially leaving undiscovered biomarkers relevant to the heterogeneity. Third, while the EPIC-Norfolk replication reproduced the direction of every association, the modest number of incident CAD cases (n = 233) limited statistical power, so most estimates did not reach conventional significance thresholds. Additional external cohorts are therefore needed to assess the generalizability of our framework across diverse populations. Fourth, circulating-protein levels can fluctuate over time, and a single measurement may not capture dynamic changes related to disease progression or treatment; future studies with repeated sampling will provide a more comprehensive view. Finally, we relied on Olink assay–based relative quantification. Results obtained with other platforms, such as the SomaScan assay, may differ, so cross-platform generalizability remains to be confirmed. Future work using clinical-grade and absolute quantification across multiple proteomic modalities will be important for eventual clinical implementation.

Overall, our findings show that baseline plasma proteomic variation can reveal structured heterogeneity among individuals who develop CAD, characterized by continuous biological gradients. The proteomic dimensions capture clinically meaningful differences, predict CAD and comorbidity risk beyond clinical variables, map CAD trajectory, and illuminate biological programs. Together, these results highlight that plasma proteomic heterogeneity can serve as a central layer bridging clinical and mechanistic heterogeneity in CAD. While further work is warranted to assess generalizability (e.g., across ancestries and across platforms) and potentially improve the protein panel, this proof-of-concept study provides a foundation for future proteomics studies aimed at refining risk stratification for individuals at high risk of developing CAD and elucidating mechanisms.

## Data Availability

UK Biobank data are accessible with an approved application (http://www.ukbiobank.ac.uk/). Individual-level EPIC-Norfolk data (https://www.epic-norfolk.org.uk/) are available from the respective investigators upon agreement.

## Code Availability

We used R v4.4.0 (https://www.r-project.org/). R packages included missForest v1.5, FNN v1.4.4.1, survival v3.7.0, monocle v2.32.0, fmsb v0.7.6, NbClust v3.0.1, furrr v0.3.1, future v1.34.0, and forestplot v0.1.0. Custom code developed for this study will be made available on GitHub upon publication, and the R script for mapping individuals with the 320 proteins onto the proteomic dimensions defined here will also be provided in the same repository.

## Acknowledgements

The authors gratefully acknowledge the participants and staff of the UK Biobank and EPIC-Norfolk studies for their important contributions to this research.

## Sources of Funding

TL has been supported by startup funding from the Office of the Vice Chancellor for Research, School of Medicine and Public Health, and Department of Population Health Sciences at the University of Wisconsin-Madison. The Yoshiji Lab is supported by the Canada Research Chairs Program (CRC-2025-00097), the Canadian Institutes of Health Research (183596), the DNA to RNA (D2R) Foundational Program, Japan Society for the Promotion of Science, and McGill University. The funders had no role in the study design, conduct, analysis, or reporting.

## Disclosures

P.N. reports research grants from Allelica, Amgen, Apple, Boston Scientific, Cleerly, Genentech / Roche, Ionis, Novartis, and Silence Therapeutics, personal fees from Allelica, Apple, AstraZeneca, Bain Capital, Blackstone Life Sciences, Bristol Myers Squibb, Creative Education Concepts, CRISPR Therapeutics, Eli Lilly & Co, Esperion Therapeutics, Foresite Capital, Foresite Labs, Genentech / Roche, GV, HeartFlow, Magnet Biomedicine, Merck, Novartis, Novo Nordisk, TenSixteen Bio, and Tourmaline Bio, equity in Bolt, Candela, Mercury, MyOme, Parameter Health, Preciseli, and TenSixteen Bio, and spousal employment at Vertex Pharmaceuticals, all unrelated to the present work. T.L. has been providing consulting services to Five Prime Sciences Inc., which was not involved in the design, execution, analysis, or interpretation of the study. S.Y. serves as an Analytical Consultant for the Broad Institute of MIT and Harvard through Precision Global Consulting. The other authors declare no conflict of interest.

## Supplemental Material

Supplementary Notes 1-4 and References 37-39,55.

Figure S1-S11

Table S1-S16

---

**Figure S1: a**, flowchart illustrating the protein selection process for incident CAD prediction. The input comprised 39,090 controls, 3,713 cases, and 2,920 proteins. The dataset was split into 70% for model development (LASSO-based selection) and 30% for model evaluation. From LASSO, 320 proteins were identified, which were then tested alongside QRISK3, PCE and PREVENT risk scores. **b**, the C-index (with 95% confidence intervals) for six Cox models: QRISK3 + LASSO, PCE + LASSO, PREVENT + LASSO, QRISK3, PCE and PREVENT. Including the 320 proteins selected by LASSO improved risk prediction beyond the baseline clinical scores alone, demonstrating the added value of proteomic data

**Figure S2: a**, dimension reduction on 320 CAD-related proteins of 3,713 incident CAD cases by PCA or **b**, UMAP; each point is an individual.

**Figure S3:** Each panel shows the top 10 diseases significantly associated with dimension 1 (upper panel) or dimension 2 (lower panel) in a Phenome-Wide Association study (PheWAS). Points indicate the regression coefficients (with 95% confidence intervals) derived from Cox regression models, interpreted as log hazard ratios for disease incidence. Conditions are listed on the y-axis, while the x-axis displays the estimated effect size with respect to dimension 1 or dimension 2. Chronic kidney disease appears most strongly associated with dimension 1, whereas metabolic disorders such as obesity and type 2 diabetes show prominent associations with both dimensions, suggesting distinct yet overlapping pathophysiological pathways captured by each proteomic dimension.

**Figure S4:** A tree plot illustrates the distribution of 233 incident CAD cases with proteomic dimensions in the validation analysis, based on 320 CAD-related proteins. Each data point represents a single individual.

**Figure S5:** For each of the 20 proteins with the largest LASSO coefficients, we performed a joint linear model (standardized protein ∼ BMI + serum creatinine) on 42,803 UK Biobank participants. The absolute Z-statistics (|Z|) for BMI and creatinine were calculated from these models. In each panel, the x-axis and y-axis represent the absolute contributions of each protein to dimension 1 and dimension 2, respectively, as derived from the DDRTree weight matrix. Points are labeled with protein names. Panels (a) and (b) show bubble areas corresponding to |Z| for BMI and creatinine, respectively. Panel (c) encodes the ratio of creatinine-to-BMI |Z| values, highlighting proteins more strongly associated with creatinine. The dotted 45° line indicates an equal contribution to both dimensions. Overall, the larger bubbles near the -axis suggest that dimension 1 captures additional renal (creatinine-related) signals, whereas dimension 2 is driven more by metabolic variation linked to BMI.

**Figure S6: a**, A tree plot illustrating the DDRTree clustering of 3,713 incident CAD cases, based on CAD-related proteins, revealing 9 distinct clusters. Each data point represents a single individual.

**b**, Radar charts depicting the mean values of nine standardized CAD-related clinical variables: body mass index (BMI), body fat percentage (BFP), hemoglobin A1c (HbA1c), alanine aminotransferase (ALT), creatinine (CREAT), triglyceride (TRIG), systolic blood pressure (SBP), high-density lipoprotein cholesterol (HDL-C) and Low-density lipoprotein (LDL) for each identified cluster. Each clinical variable was standardized with rank-based inverse-normal transformation within the 3,713 incident CAD cases. The dotted line represents the mean value of zero.

**Figure S7:** Radar charts depicting the mean values of nine standardized CAD-related clinical variables: body mass index (BMI), body fat percentage (BFP), hemoglobin A1c (HbA1c), alanine aminotransferase (ALT), creatinine (CREAT), triglyceride (TRIG), systolic blood pressure (SBP), high-density lipoprotein cholesterol (HDL-C) and Low-density lipoprotein (LDL) for each identified cluster after adjustment for statin, aspirin and antihypertensive use. The dotted line represents a mean value of zero.

**Figure S8:** Violin plot showed the kernel-density distributions of dimension 1 and dimension 2 for incident CAD cases presented without MI (grey) and with MI (green). Within each violin, the white box marks the inter-quartile range (IQR) and the black horizontal line indicates the median; whiskers extend to 1.5 × IQR.

**Figure S9:** Grouped bar charts illustrate the proportion of participants who subsequently developed incident type 2 diabetes (T2D), obesity, hypertension, chronic kidney disease (CKD), or heart failure (HF) across the nine coronary artery disease (CAD) clusters, within 10-year of proteomic measurement. Each cluster on the x-axis includes five bars (one per condition), and the y-axis represents the corresponding proportion of incident cases. Across the nine CAD clusters, participant counts were Cluster 1 = 723, Cluster 2 = 555, Cluster 3 = 107, Cluster 4 = 490, Cluster 5 = 159, Cluster 6 = 575, Cluster 7 = 251, Cluster 8 = 271, Cluster 9 = 582.

**Figure S10:** Cox proportional hazard models were built between each disease and each clinically adjusted proteomic dimension from all 42,803 participants. **a**, association for clinically adjusted dimension 1, upper triangles indicate a positive association between the clinically adjusted proteomic dimension values and disease risk, whereas lower triangles denote a negative association. Diseases were considered statistically significant associated with clinically adjusted proteomic dimensions if their FDR-adjusted p-value was below 0.01. **b**, association for clinically adjusted proteomic dimension 2. **c**, combined comparison of disease associations across the two clinically adjusted dimensions. Each point represents a disease endpoint, positioned by its signed −log_10_(P) for clinically adjusted dimension 1 (x-axis) and clinically adjusted dimension 2 (y-axis). The related signs were determined by the direction of associations.

**Figure S11:** Regression coefficients with 95% confidence intervals (x-axis) are derived from linear regressions that assess the associations between each clinical variable (rows on the y-axis) and each clinically adjusted proteomic dimension (clinically adjusted dimension 1 in blue or dimension 2 in purple). The dashed vertical line at zero indicates no effect. Abbreviations for clinical variables include WBC_count (White blood cell count), NEU_count (Neutrophil count), VAT (Visceral adipose tissue), glucose (Blood glucose), GGT (Gamma-glutamyl transferase), AST (Aspartate aminotransferase; AST), CysC (Cystatin C), urea (Urea), UM (Urine microalbumin), UCE(Urine creatinine, enzymatic assay), KPV (Total kidney parenchyma volume), MKV (Mean kidney volume).

## Notes

### Author Declarations

The North West Multi-centre Research Ethics Committee of the United Kingdom National Health Service gave ethical approval for the UK Biobank component of this work. The Norfolk Research Ethics Committee of the United Kingdom National Health Service gave ethical approval for the EPIC-Norfolk component of this work. The present analysis was approved by the UK Biobank (application no. 73958) and the Norfolk Research Ethics Committee (no. 05/Q0101/191). All participants provided written informed consent.

